# Treatment of Spontaneous Subarachnoid Hemorrhage: A 20-year National Inpatient Sample Review

**DOI:** 10.1101/2024.05.01.24306726

**Authors:** Joshua Feler, Dylan Wolman, Radmehr Torabi, Jia-Shu Chen, Abigail Teshome, Rahul Sastry, Curtis Doberstein, Mahesh Jayaraman, Krisztina Moldovan, Ali Mahta

## Abstract

**Background:** Aneurysmal subarachnoid hemorrhage is a major source of morbidity and mortality, and its management has undergone foundational changes over the last 2 decades. We reviewed the National Inpatient Sample (NIS) to outline the changes in severity of illness, surgical management, and patient outcomes over time.

**Methods:** A retrospective cohort of admissions for spontaneous subarachnoid hemorrhage (SAH) in the NIS from 2001-2020 was reviewed, including those that underwent microsurgical or endovascular surgery to secure a ruptured aneurysm. National incidence was calculated, and multivariable regression was used to identify changes in incidence and outcome through time, segmented by epoch.

**Results:** Review of the NIS identified 450,723 SAH patients, of which 182,151 underwent surgical aneurysm treatment. The incidence of spontaneous SAH fell −0.127 per 100,000 person-years each year [95%CI: −0.164, −0.0887]). Among patients surgically treated for aneurysmal SAH, the proportion of patients <50 years old fell from 40% to 30% between first and final epochs, and the proportion of those in the lowest stroke scale category, roughly equivalent to Hunt & Hess grade 1 or 2, fell from 68% to 49%. The proportion treated by microsurgery fell from 70% to 23% in favor of endovascular surgery. Hospital mortality among these treated cases was stable at 13% throughout the study period despite increasing illness severity indices. After adjustment, there was a 41% reduction of odds of hospital mortality in the final epoch compared with the first.

**Conclusions:** The incidence of hospitalization for spontaneous SAH fell between 2001 and 2020. Patients undergoing surgery to secure an aneurysm were more severely ill through time yet experienced a stable hospital mortality rate.

## Background

The two decades from 2001 to 2020 saw numerous changes in the medical and surgical management of patients presenting with aneurysmal subarachnoid hemorrhage (aSAH). This period saw a randomized trial of prophylactic triple-H (hypertension, hypervolemia, hemodilution) therapy fail to show benefit in improving cerebral blood flow,^1^ growing evidence against the routine use of anti-fibrinolytic therapy,^2,3^ and the publication of the ISAT^4^ and BRAT^5^ trials comparing the efficacy of microsurgical and endovascular surgeries for ruptured aneurysms.^6^ These changes to aSAH management occurred alongside an evolving demographic backdrop, with an aging and increasingly diverse population in the United States.^7^ Furthermore, the percentage of aSAH patients presenting with poor clinical grade has increased over the last two decades, which has important implications for the efficacy of each intervention strategy.

The National Inpatient Sample (NIS), a nationwide all-payer inpatient database containing a probability sample of discharges, has been used for many purposes including demonstrating the volume/outcome relationship,^8^ reporting on rare complications such as acute lung injury,^9^ and identifying practice pattern variations in tracheostomy.^10^ An estimating function for Hunt and Hess grade has also validated to improve clinical specificity.^11^ However, many of these analyses conclude at the International Classification of Disease (ICD), ninth edition (ICD9) to ICD, tenth edition (ICD10) transition in 2015 as coding changes require the accumulation of several years of data to re-evaluate, especially as early evaluations of aSAH suggested a possible bias towards differential case capture.^12^

However, recent data accumulation has allowed exploration of the ICD10 period. Prior analyses of the NIS have explored the decreasing relative risk of admission due to aSAH compared with admission for elective securement of an unruptured aneurysm or against other unrelated causes for hospitalization to suggest a decreasing relative risk of admission with aSAH through time.^13,14^ This format, though useful for understanding the evolution of risk factors and practice patterns through time, obscures the total burden of disease in the setting of rising numbers of elective treatments. We aim to explore the evolution of patient severity of illness and outcomes through re-analysis of a larger set of admissions in the NIS from 2001 to 2020.

## Methods

### Data source and inclusion criteria

Data from the NIS from 2001-2020 were included. Admissions were identified by the presence of ICD9 and 10 codes consistent with non-traumatic SAH in the primary diagnosis position. Records were included with ages ≥18 years, non-elective admission type. Cases with diagnoses for other non-aneurysmal vascular malformations like arteriovenous malformations or diagnoses for head trauma were excluded. Treated aneurysmal SAH were further identified by the presence of procedure codes for endovascular or microsurgical procedures to secure an aneurysm. ICD code dictionaries are included in Supplementary Material 1 and were modeled after the schema previously described by Chang *et al*..^15^ Annualized population estimates for each age group were collected from intercensal estimates by the United States Census Bureau.^16^ This project was determined to be exempt from IRB review (Lifespan IRB #1466665)

### Measures and outcomes

Patient were stratified into those less than 50 years old and then into decades thereafter until reaching a category for patients greater than 80 years of age. NIS SAH Severity Score (NIS-SSS) was calculated for each patient and is divided into groups I, II, and III that roughly correspond to Hunt & Hess Grade (HH) 1-2, HH3, and HH4-5. This model was initially validated using ICD9 code definitions but has been previously adapted for use with ICD10 codes.^13^ Elixhauser comorbidity indices (ECI), a comorbidity scale capturing 30 comorbidities and weighted as described by van Walraven *et al*.,^17^ were calculated from ICD9 and 10 diagnosis codes. All Patient Refined-Diagnosis Related Group (APRDRG) illness severity and mortality risk indices, disease specific abstractions that measure a patient’s resource intensity according to both primary and secondary diagnoses, were also captured. Procedural proxies for illness severity including CSF diversion, ventilation, and early ventilation (<72h after admission) were identified. Non-home discharge included discharge to institutionalized settings and mortalities. Tracheostomy and gastrostomy procedures were identified as outcome measures. Data years were divided into 4 epochs spanning 2001-2005, 2006-2010, 2011-2015, and 2016-2020, defined inclusively. Incidence of the disease process was calculated as estimated admissions within an age group divided by the national population within that age group. The NIS is a nationally maintained, anonymized dataset, and this analysis was exempt from Institutional Review Board review. Data will be made available upon reasonable request to the corresponding author.

### Effects of ICD9 to ICD10 transition

The ICD9 to ICD10 transition was evaluated with an interrupted time series analysis for selected variables. The transition from ICD9 to ICD 10 occurred in 2015. Admission volumes and selected admission level covariates were modeled with the function:

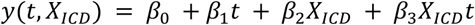

where *y* is the value being modeled through time, *β*_*n*_ are model parameters, *t* is the time since study initiation, and *X*_*ICD*_ is a dummy variable set to 0 pre-transition to ICD10 and set to 1 after transition to ICD10. In this model, the presence of *β*_2_ or *β*_3_ values that are significantly different from 0 suggests a change associated with the coding system, whereas a *β*_1_value significantly different from 0 suggests a linear change through time independent of the ICD version. We performed a sensitivity analysis around selection of cutoff years for ICD9 to ICD10 to test the robustness of findings of data discontinuity associated with this change. Sensitivity analysis was also performed in multivariable modeling by calculating the primary models with and without the final epoch (which corresponds to the ICD10 period) and by calculating the models with the addition of an independent variable coding ICD type.

### Statistics and Outcome Modeling

National estimates were produced after applying NIS weighting and accounting for the stratified survey design of the sample. All comparisons were performed considering survey weights. Least squares regression weighted by the inverse of variance was used to evaluate trends in annualized counts. Weighted logistic regression was used to evaluate for associations between hospitalization-level characteristics and primary surgical treatment modality (microsurgery or endovascular surgery for aneurysmal occlusion), hospital mortality, and non-home discharge. Model parameters were selected based on results from prior studies and included age category, sex, NIS-SSS grade, surgical treatment modality, hospital size, and hospital region. Hospitalizations in which both microsurgery and endovascular surgery for aneurysm securement were employed were excluded from multivariate analysis. There were many missing data points for race and zip code income quartile disproportionately impacting the first epoch, so a primary model was calculated without these features and a secondary model was calculated with these features included. NIS-SSS was used in the primary models, though they were also calculated with a procedure-based SSS to test the assumption that there may exist changes in coding practices that might lead to upcoding of otherwise similar patients through time.^18^ Models were compared with Akaike Information Criterion (AIC) if not nested and with analysis of variance (ANOVA) if nested. Collinearity was assessed by variance inflation factors (VIF). The reference epoch to which the latter epochs were compared was 2001-2005. P-values were adjusted with Holm’s method to yield a q-value. Significance in these models was defined as q-value <0.05. All statistics were calculated in RStudio (Version 2023.09.1+494).

## Results

### Total SAH population

An estimated total 450,723 admissions with spontaneous SAH were identified in the study period with an incidence of 9.56 per 100,000 person-years. Through time, the total admission count non-significantly decreased (−74.4/year [95%CI: −165, - 17.3], p=0.10). The incidence decreased −0.127 per 100,000 person-years per year (95%CI: −0.164, −0.0887) to a minimum of 8.07 per 100,000 person-years in 2020. Changes in counts and incidence among age groups for spontaneous SAH are plotted in **Figure 1**. Total admissions decreased within the <50-year-old group (−142/year [95%CI −176– −107]) while those in the 60-70 and 70-80 groups increased (72.0/year [95%CI 54.0-90.2], 28.4/year [95%CI 11.1-45.7]). The incidence of spontaneous SAH decreased for all age groups. Across the whole study period, the mortality rate was 24.5%, with the odds of mortality decreasing by 1.44%/year (95%CI 1.17%–1.71%). The ICD coding version was not a significant parameter in interrupted time series regression for total counts, incidence, or odds of treatment.

**Figure 1:**
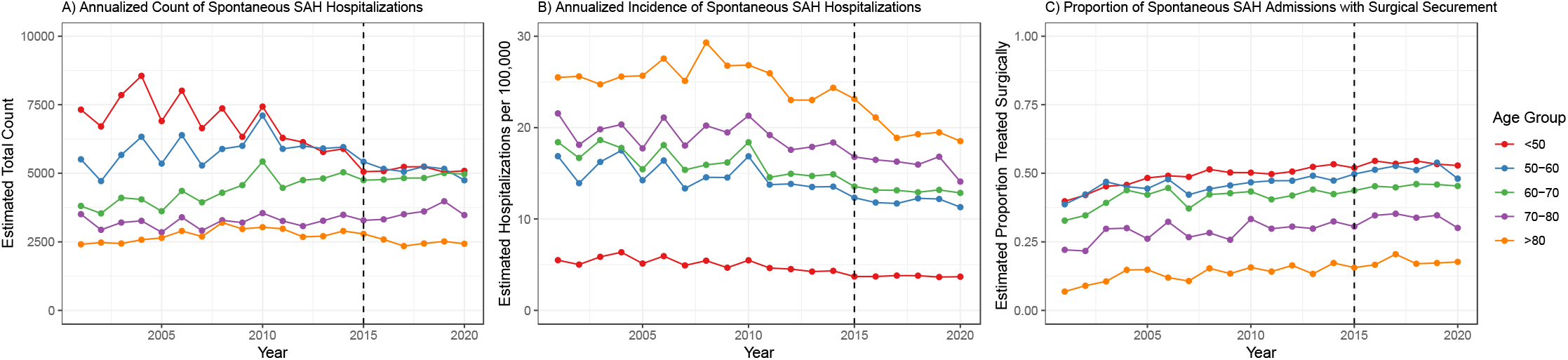
A) Total admissions for spontaneous SAH. B) Incidence of hospitalization for spontaneous SAH. C) Proportion of surgically treated aSAH.

### Surgically Treated aSAH Population

A total of 182,151 (40.4%) patients underwent surgical aneurysm treatment (**Table 1**). The odds of surgical treatment for an aneurysm increased through time at a rate of 1.85%/year [95%CI 1.41%-2.29%], yielding an increased count of hospitalizations of 77.19/year (95%CI: 22.1–132) with an unchanged incidence. Increased hospitalizations were primarily among older adults and those in NIH-SSS groups II and III (**Figure 2**). The age group <50 had decreasing total admissions (−33.3/year [95%CI: −45.5– −12.9]); 50-60 were unchanged; and higher age groups increased (60-70: 54.2/year [95%CI: 43.9, 64.4], 70-80: 26.5/year [95%CI: 17.4–35.6], >80: 13.0/year [95%CI: 8.39-17.5]). When divided by age group, incidence changed only for those <50 years old, among whom it decreased through time (−0.0285 per 100,000 person-years per year [95%CI −0.0444– −0.0133]). Admissions of NIS-SSS grade I decreased through time (−75.8/year [95%CI: −109, −42.9]) while those in grade II and III increased (39.3/year [95%CI: 21.8– 56.7]; 125/year [95%CI: 112–139]).

**Table 1:**
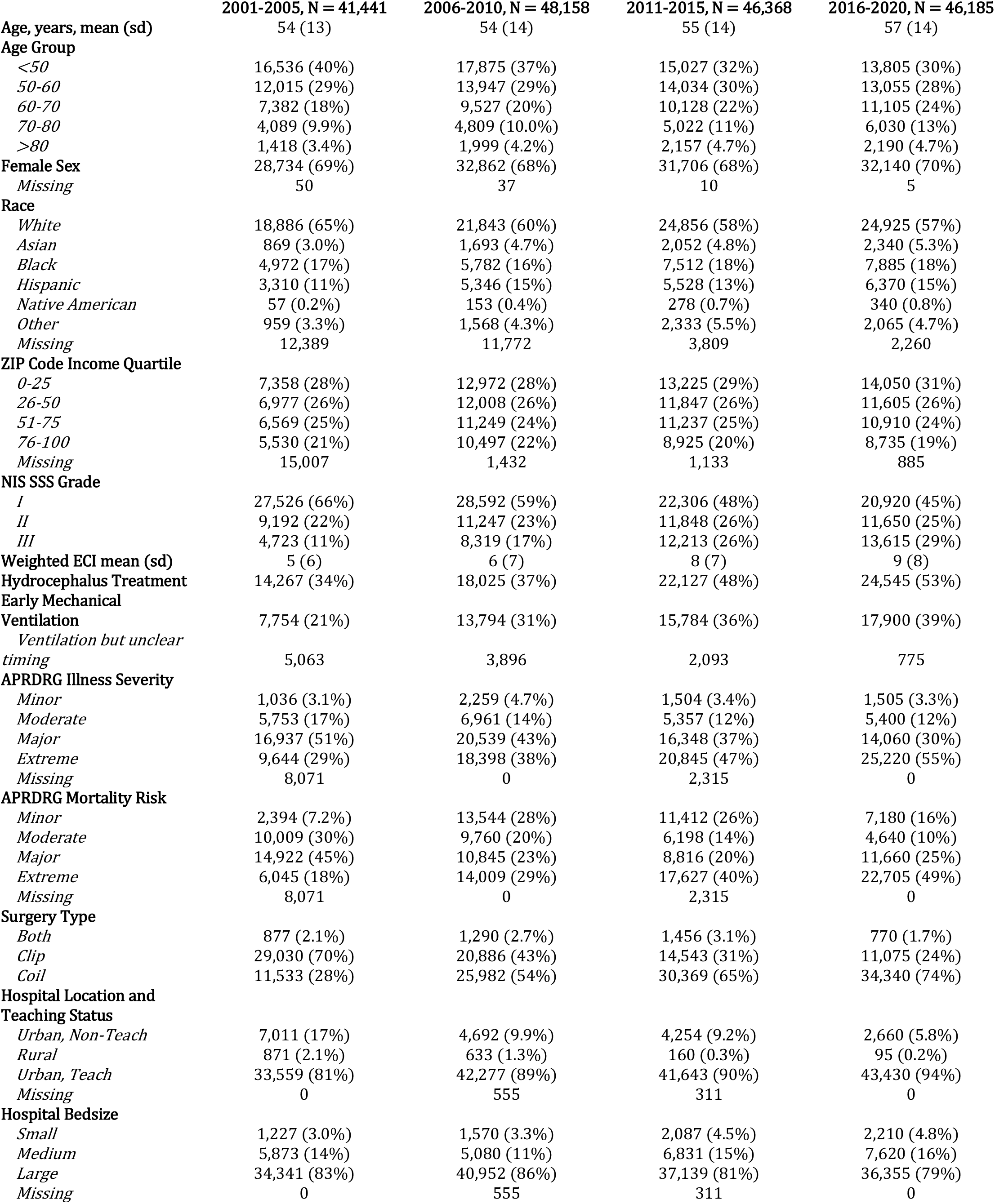
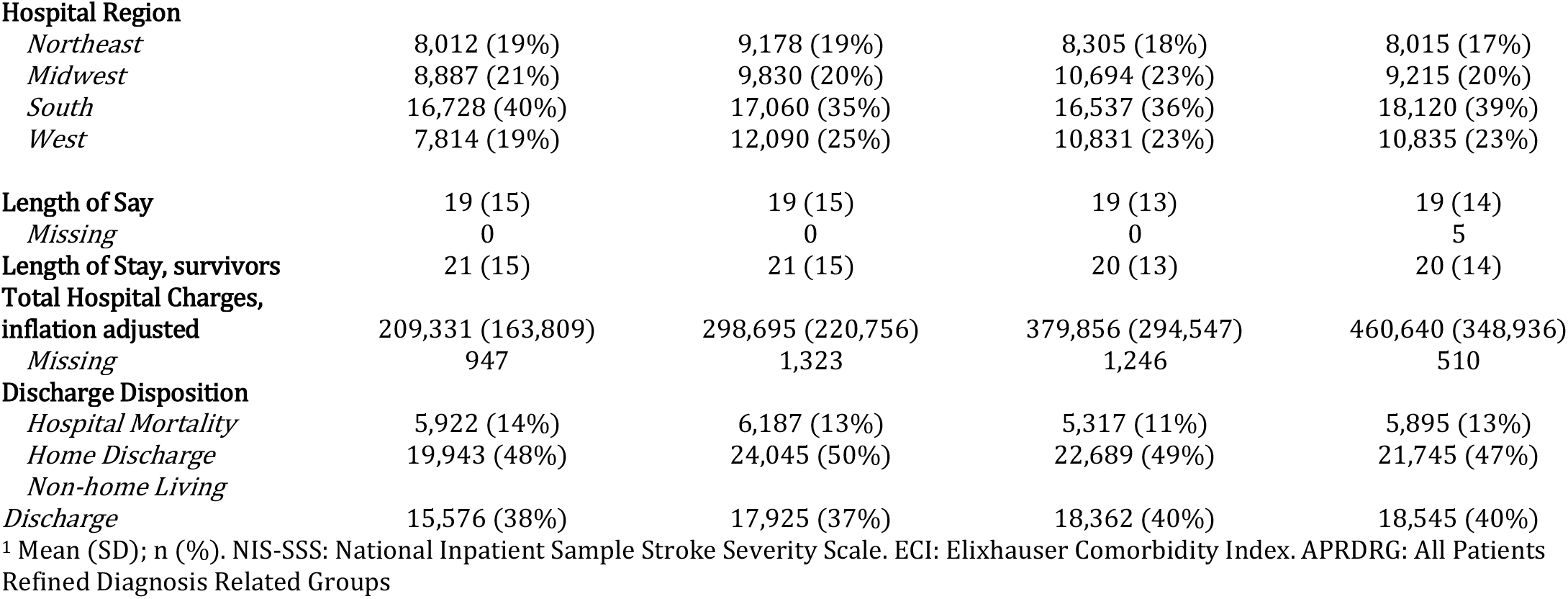
Characteristics of surgically trated aSAH patients stratified by epoch.

**Figure 2:**
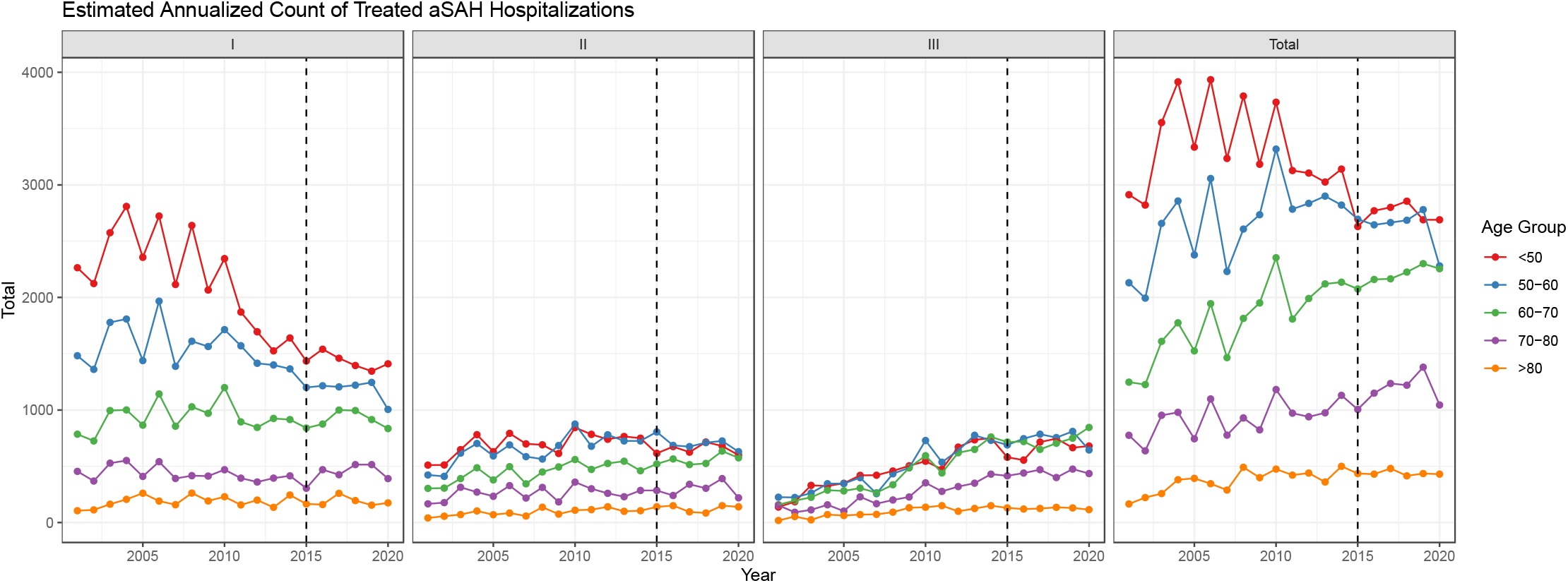
Annualized estimates of admission counts of treated aSAH by age group and NIS-SSS grade.

Mean age increased from 54 to 57 years between first and final epoch (p<0.001), and the distribution of age groups changed across the study period with patients age <50 years old falling from 40% to 30% between the first and final epochs (p<0.001). The proportion of admissions with non-white races increased between the first and last epoch (65% vs. 57%, p<0.001). The distribution of income quartiles was skewed towards lower income quartiles in all epochs, and the skew worsened slightly through time with increased admissions in the lower half of income quartiles from 54% to 57% between first and final epochs.

Indices for baseline and presentation illness severity including NIS-SSS, ECI, APRDRG illness severity, APRDRG mortality risk, hydrocephalus treatments, and early ventilation all increased between the first and last epoch and are plotted in **Figure 3**. Notably, the proportion of NIS-SSS grade I hospitalizations fell from 66% to 45% (p<0.001). Treatments for hydrocephalus rose from 34% to 53% (p<0.001) while early mechanical ventilation rose from 30% to 39% (p<0.001). The early mechanical ventilation rate is repotted between the second and final epochs given the missingness of procedure timing in the first epoch. Through time, hospitalizations were concentrated in urban teaching hospitals, rising from 81% to 94%, and there was a small shift away from the largest hospitals, declining from 83% to 79% (p<0.001).

**Figure 3:**
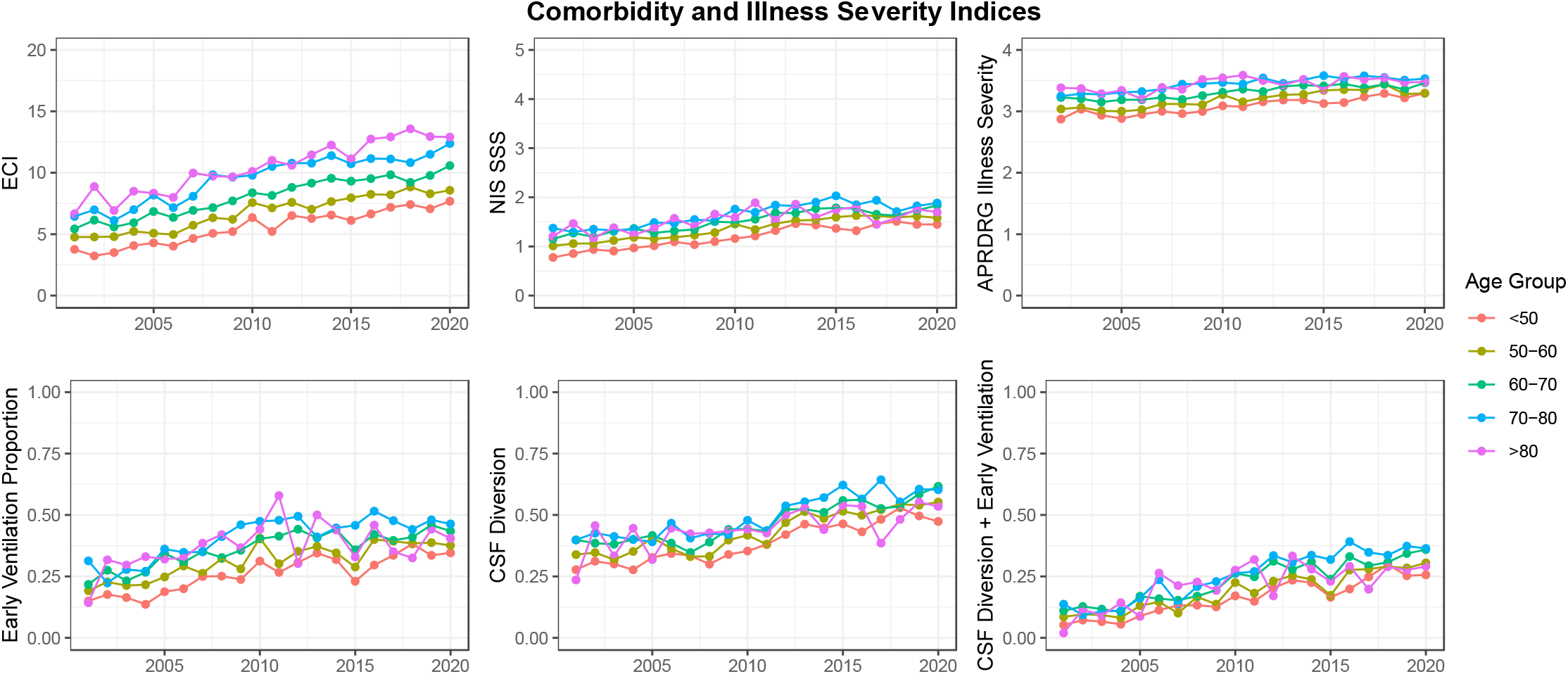
Annualized estimates of comorbidity indices by age.

#### ICD9 to ICD10 Transition

In ordinary least squares regression for admission counts and linear regression for age, surgical modality, and ECI, model parameters associated with changes to ICD10 coding were non-significant consistent with an absence of evidence of data discontinuity for these features. In the model for NIS-SSS score proportion, the parameter associated with ICD coding system change was significantly different than 0 (β_2_ = 0.16 [-0.00250–0.323], β_3_ = −0.0442 [95%CI: −0.0721–-0.0162]) suggesting an under-estimation of NIS-SSS grade in the ICD10 epoch relative to the ICD9 epoch.

### Changes in Surgical Modality through Time

From the first to final epochs, the proportion of hospitalizations in which microsurgery was performed decreased from 70% to 23% (p<0.001). This change occurred over all age and NIS-SSS groups (**Figure 4**). This effect was confirmed in unadjusted and adjusted models, with the odds of undergoing microsurgery decreasing in all the latter epochs to a minimum of adjusted odds of 0.13 (95%CI: 0.11, 0.15) between the final and first epochs.

**Figure 4:**
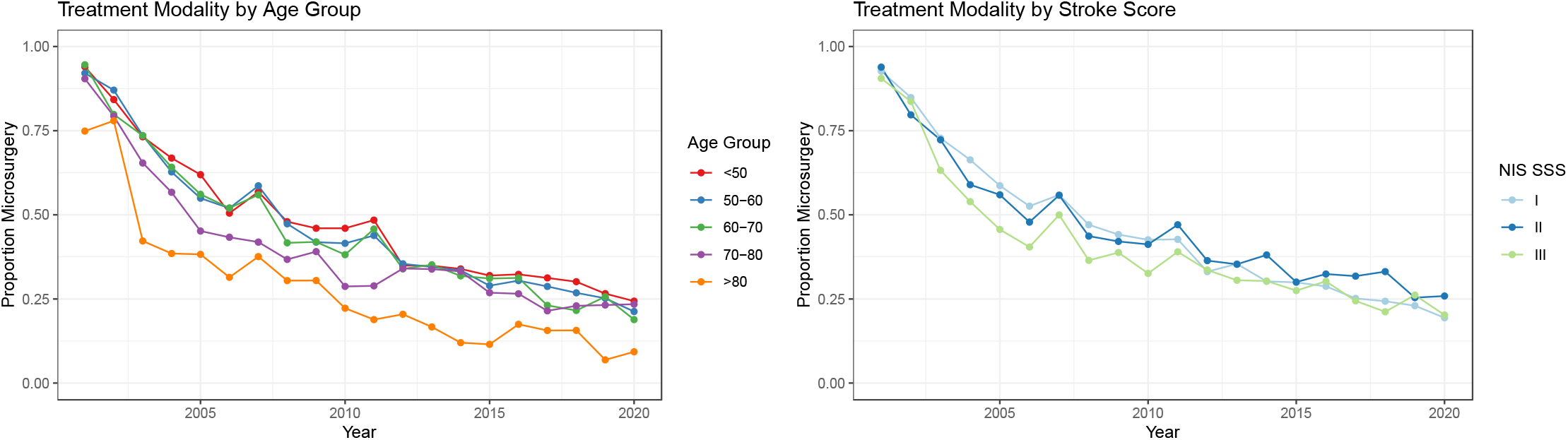
Annualized estimates of surgical treatment modality by age group and NIS SSS grade.

In the unadjusted model, older age groups exhibited decreasing odds of undergoing microsurgery, with a minimum among those >80 years old (OR0.35 [95%CI: 0.31, 0.40]). Compared with NIS-SSS grade I, both NIS-SSS grade II and III were associated with decreased odds of undergoing microsurgery (OR 0.86 [95%CI: 0.81, 0.92] and OR 0.57 [95%CI: 0.53, 0.61], respectively). Compared with the urban non-teaching hospitals, hospitalizations at urban teaching hospitals had decreased odds of microsurgery (OR 0.63 [95%CI: 0.55, 0.73]), while those admitted to rural hospitals were not different. The odds of microsurgery did not vary across hospital regions.

In the adjusted model, the association between age groups remained with similar magnitude. Hospital location and teaching status was no longer significantly associated with microsurgical clipping. Compared with NIS-SSS grade I, NIS-SSS grade III remained associated with decreased odds of microsurgery (aOR 0.79 [95%CI: 0.73, 0.84]) but NIS-SSS grade II was not significantly different than grade I. After adjustment, Western hospital region was associated with increased odds of microsurgery (aOR:1.29 [95% CI 1.09, 1.52]) compared with the Northeast. All model parameters are given in supplementary table 3.

### Predictors of Mortality

The proportion of hospital mortalities was 13% over the entire study period and significantly but not substantially decreased between first and final epoch (14% vs. 13%). Annualized point estimates for hospital mortality proportions by age group and NIS-SSS are plotted in Figure 4 showing progression of mortality rates with increasing age and increasing NIS-SSS grade. Without adjustment, there were only decreases in the odds of hospital mortality through time seen in the 2011-2015 epoch (0.77 [95%CI: 0.69, 0.85]), without significant changes in the other epochs. Progressively increased odds of hospital mortality were associated with increasing age category peaking at OR 3.90 [95%CI: 3.44, 4.42] for the group greater than 80 years old, with increasing NIS-SSS category (NIS-SSS grade II OR 2.94 [95%CI: 2.70, 3.19], NIS-SSS grade III OR 4.35 [95%CI: 4.00, 4.73]), and with increasing ECI (OR 1.03 [95%CI: 1.03, 1.03] per unit increase). Decreased odds of hospital mortality were associated with urban teaching hospitals (OR 0.69 [95%CI: 0.64, 0.77]).

In the multivariable model, decrements in the odds of mortality were more apparent with all epochs associated with decreased adjusted odds of mortality (2006-2010 aOR 0.75 [95%CI: 0.67, 0.84], 2011-2015 aOR 0.53 [95%CI: 0.48, 0.60], 2016-2020 aOR 0.58 [95%CI: 0.52, 0.65]. Female sex was associated with a reduced hospital mortality (aOR 0.89 [95%CI: 0.83, 0.96]). Other parameters were overall similar to the unadjusted model. Treatment modality was not associated with hospital mortality in unadjusted or adjusted analyses. All model parameters are given in supplementary table 4.

Alternative models were calculated using mechanical ventilation and hydrocephalus treatments in place of the NIS-SSS given the possible data discontinuity and lead to no major changes in parameter estimates. Exclusion of the final epoch, most associated with the ICD10 period, also did not meaningfully change model estimates. ANOVA testing showed that the addition of a binary ICD coding system term did not improve model fit in prediction mortality or non-home discharge.

### Hospital Outcomes

Mean length of stay among survivors decreased minimally with an average of 21 days in the first epoch and 20 in the last. The proportion of non-home living discharges rose subtly between first and final epoch (38% vs. 40%, p< 0.001). Average hospital charges more than doubled between the first and final epochs ($209,331 vs. $460,640).

### Race and Income Models

There were many records missing socioeconomic data, especially in the first epoch. For example, 30% of records in the initial epoch lacked data for race compared with 5% in the final epoch. The models above were recalculated with income quartile and race, though given the pattern of data missingness, these findings more reflect the latter part of the study period.

Regarding surgical modality for aneurysm securement, in adjusted analysis Hispanic race was associated with microsurgery (OR 1.16 [95%CI: 1.06, 1.28]). In adjusted analysis, this finding remained and the other race category was also associated with increased odds of microsurgery (aOR 1.33 [95%CI: 1.13, 1.57]). With respect to income, there were no differences in surgical modality.

Black and Hispanic races were associated with decreased unadjusted odds of mortality (OR 0.63 [95%CI: 0.57, 0.70] and OR 0.80 [95%CI: 0.72, 0.90], respectively), the former of which was preserved after adjustment but the latter was not. Without adjustment, income quartile was not associated with mortality, but after adjustment the highest income quartile was associated with decreased odds of hospital mortality (aOR 0.83 [95%CI: 0.74, 0.93])

## Discussion

In this study, we characterized longitudinal changes to the incidence, surgical management, and hospital outcomes of patients with spontaneous SAH across two decades in the NIS. Although the incidence of spontaneous SAH decreased, the proportion of those undergoing surgical aneurysm treatment increased through time with an otherwise stable incidence of surgically treated aSAH. Among aSAH patients, the proportion who underwent microsurgical aneurysm treatment declined, but a greater proportion of older and more severely ill patients were treated with notably preserved hospital mortality rates. Overall, these data suggest that surgically treated aSAH is increasingly a disease of older adults and that there are relatively improved outcomes with the national standard of care after presentation with aSAH.

### The incidence of SAH and treatment modality

Our results suggest a decreasing incidence of spontaneous SAH through time, with an average incidence of 9.8 hospitalizations per 100,000 person-years for the study period. Notably, this definition includes heterogeneous disease processes with known different outcome profiles (e.g. perimesencephalic SAH) and excludes deaths prior to hospital admission. The latter, an important contributor to the incidence of SAH, is difficult to quantify given that those who do not survive to admission may not receive a diagnosis. Prior reports from Minnesota and Finland suggest a spontaneous SAH incidence ranging from 12-26%, while a US-based study including autopsy diagnosis suggested a stable aSAH incidence from 2007-2017 with a mean 11.4 cases per 100,000 person-years, but this study lacked the volume of cases represented in our dataset.^19–21^ Prior analyses of the NIS from 2004-2014 have attempted to describe the relative incidence of hospitalization for aSAH by comparing with hospitalizations for elective treatment of aneurysms or other disease processes to suggest a decreasing risk of admission for aSAH, but each has been methodologically limited in that they reference non-fixed indices and have structural selection biases intrinsic to the NIS.^13,14^

The selection of admissions including surgical aneurysm treatment was intended to select the population of patients with presumed survivable aSAH. There was an overall increased incidence of hospitalizations for treated aSAH with increasing representation of patients with advanced age and greater illness severity through time, similar to what has been reported in other NIS studies of aSAH and concordant with a prior single-center study demonstrating a 7.2% increase in high Hunt & Hess grade admissions over 30 years.^13,14,22,23^ Although the interrupted time series regression of NIS-SSS suggested a possible data discontinuity, the ICD10 period was characterized by relative underestimation which, if corrected, would further enrich the final epoch with high grade hospitalizations and make this trend even more pronounced.

The effect of medical optimization and elective aneurysm treatment on SAH incidence is a matter of public health importance. Although our analysis did not reveal a decrease in the incidence of surgically treated aSAH through time, beneficial effects may be obscured by changes in admission practices. We found an increase in the number of hospitalizations for aSAH over time, particularly among older and more severely ill patients, and we hypothesize that improved pre-hospital care may lead to fewer pre-admission mortalities while microsurgical and endovascular advancements may result in more optimistic and aggressive aneurysm treatment despite advanced age or severe neurologic impairment.

Our analysis demonstrated a striking trend favoring endovascular surgical aneurysm management, increasing from 23% of hospitalizations in 2001-2005 to 70% of hospitalizations in 2016-2020. The evolving treatment pattern broadly reflected changes to national guidelines through the study period, from “can be beneficial” in 2009^24^ to “superior” with respect to neurological outcomes in 2023^6^ – published after the end of the study period but reflecting practice paradigms that developed therein. However, we found a regionalized paradigm with increased odds of microsurgery among hospitalizations in the Western region, consistent with a prior Vizient database analysis which additionally noted increased adjusted odds of poor outcomes associated with the Western region which were not duplicated in our own analysis.^25^ The balance of treatments in this region still favors endovascular, but to a lesser extent than in the comparator, the northeast region.

The growing prominence of older adults with aSAH between 60 and 80 years of age is consistent with the general aging of the population of the United States.^7^ Similarly, the declining proportion of white patients reflects national changes in demography, as the proportion of white United States citizens fell through the study period from 69% to 58%.^16^ The slight bias of increasing aSAH within the lower income demographic is consistent with increased prevalence of stroke risk factors among patients of lower socioeconomic status and a potential lack of access to or desire for elective treatment.^26^ In conjunction with the decreased odds of mortality in the highest income group this raises concern for a widening health disparity, as further discussed by Schupper *et al*.^27^

### Improving outcome profiles

The unadjusted hospital mortality improved for spontaneous SAH over time but not surgically treated aSAH, with a stable proportion of 13% through the study period. However, it is notable that mortality was unchanged despite increasing proportions of hospitalizations for older patients and those with severe disease. There was a clear improvement in adjusted odds of mortality through time that may be influenced by improved case selection among high-grade patients and advances in medical and surgical care. However, this finding must be approached with cautious optimism, as at the cohort level in all epochs, the largest disposition category was to an institutional setting. Many of these patients may improve through time. A prior study found that among aSAH patient discharges to institutionalized settings, 47.5% are to an acute rehabilitation facility and 52.4% are to long-term care, with 41% of the latter group improving to mRS <3 by 90-days.^28^ A Netherlands-based study found that 38% of patients discharged to skilled nursing facilities progressed to a lower level of care.^29^ Similarly, in the long term follow up of the BRAT study, a mRS of >2 was noted in 67% at discharge and 30% at 1-year,^30^ while a Spanish analysis found that the 3-month proportion of patients with poor neurologic status was 42%, declining to 36% at 1- and 5-years.^31^ Taken together, these data suggest that a substantial proportion of the non-home, living discharges may ultimately have favorable neurological outcomes.

### The impact and limitations of case selection through time

A major limitation of assessing the impact of hospitalization-level variables is that selection biases may evolve through time with ambiguous impacts on outcomes modeling. As such, the impact of aneurysm treatment method–microsurgical or endovascular–should be interpreted extremely cautiously as the types of patients selected for either modality have evolved through time along dimensions that may not be captured by this dataset, including anatomic considerations, the presence of intraparenchymal hemorrhage, provider expertise, resource availability, or even patient or family preference. Without granular patient-level anatomic data or records of need for re-treatment or treatment-related complications, assertions about the impact of procedure choice on hospital outcomes are extremely limited. While we found decreased unadjusted odds of mortality and increased proportion of patients treated by endovascular surgery across all epochs, it is important to not interpret this association as a causal relationship. Rather, these data suggest that the overall paradigm of care improved over time.

### Limitations

With respect to neurologic injury, the NIS has known deficiencies. Of greatest import, outcomes assessments other than mortality are limited, especially when considering the proportion of patients with delayed neurological recovery. Hospitalizations are codified by ICD codes that are known to have imperfect sensitivity and specificity,^32,33^ and these values may change unpredictably through time^18^ with possible upcoding of comorbidities and disease modifiers to maximize reimbursement by patient complexity.^34^ Other authors have previously evaluated the effect of the transition on prevalence of neurological disease and noted a 20 patient per month increase in prevalence of SAH after transition.^12^ We did not observe this effect perhaps due to increased years of data and annual rather than monthly analysis. These ICD coding related effects were mitigated by calculating epoch-specific models during which coding is likely more homogenous. Also, we utilized calculation of separate models that depended only on procedures that mark severe disease, which may be less sensitivity to code drift. This study also excludes relevant subpopulations such as perimesencephalic SAH as they do not undergo surgeries, though the outcome profile for this population is distinct.

## Conclusions

Our analysis of the NIS from 2001-2020 suggests that a greater number of patients with aSAH survived to hospital arrival and were selected for further care, with an increasing proportion of older and more severely ill patients. Despite this change, the standard of care improved to deliver decreased rates of hospital mortality. While there was a profound rise in prevalence of endovascular surgical aneurysm treatment, there was no clear impact on hospital mortality based on treatment modality. The true national incidence of aSAH remains unclear, however the standard of care appears to be improving, with superior outcomes through time for those that survive to hospital arrival and are admitted for further care.

## Data Availability

Research data and statistical code will be made available upon request.

## Acknowledgments

We would like to thank Dr. Janine Moline PhD MS for her guidance in formulating the statistical analysis described herein.

## Supplemental Tables

**Supplemental Table 1:**
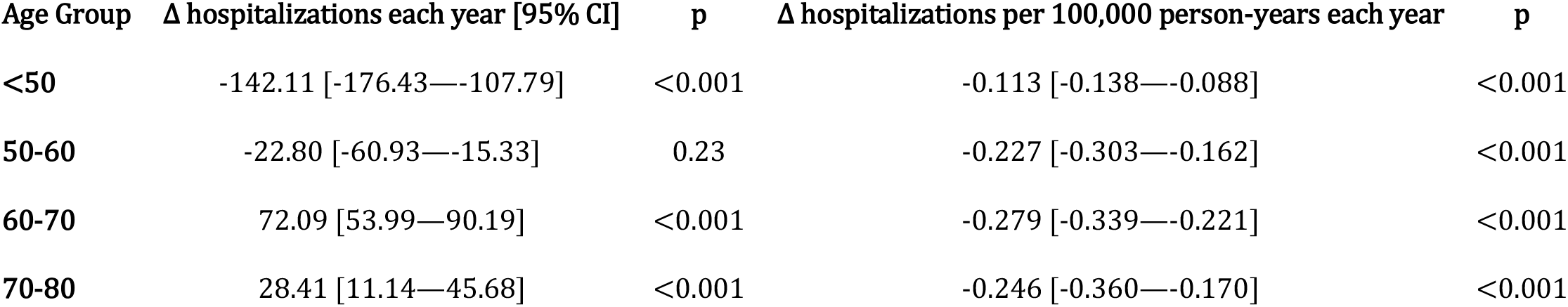
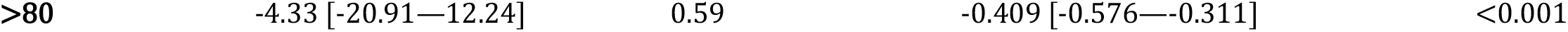
Linear regression of count and crude rates of spontaneous subarachnoid hemorrhage hospitalizations.

**Supplemental Table 2:** Linear regression of count and crude rates of surgically treated aneurysmal subarachnoid hemorrhage hospitalizations.

| Age Group | Δ hospitalizations each year [95% CI] | p | Δ hospitalizations per 100,000 person-years each year | p |
| --- | --- | --- | --- | --- |
| <50 | -33.3 [-54.5— -12.0] | 0.0 | -0.029 [-0.044— -0.013] | 0.001 |
| 50-60 | 19.9 [-1.2—41.0] | 0.1 | -0.029 [-0.74—0.007] | 0.095 |
| 60-70 | 54.2 [43.9—64.4] | <0.001 | -0.027 [-0.075— -0.000] | 0.051 |
| 70-80 | 26.5 [17.4—35.6] | <0.001 | 0.032 [-0.38—0.76] | 0.490 |
| >80 | 13.0 [8.4—17.4] | <0.001 | 0.068 [0.14—0.100] | 0.012 |

**Supplemental Table 3:** Model parameters for multivariable regression for microsurgical treatment.

|  | Unadjusted Model |  |  |  | Adjusted Model |  |  |  |  |
| --- | --- | --- | --- | --- | --- | --- | --- | --- | --- |
|  | OR<sup>1</sup> | 95% CI<sup>1</sup> | p-value | q-value<sup>2</sup> | OR<sup>1</sup> | 95% CI<sup>1</sup> | p-value | q-value<sup>2</sup> | VIF<sup>1</sup> |
| Female Sex | 1.03 | 0.99, 1.08 | 0.2 | >0.9 | 1.09 | 1.04, 1.14 | <0.001 | 0.007 | 1.0 |
| Age Group |  |  |  |  |  |  |  |  | 1.1 |
| <50 | — | — |  |  | — | — |  |  |  |
| 50-60 | 0.86 | 0.81, 0.91 | <0.001 | <0.001 | 0.91 | 0.86, 0.96 | <0.001 | 0.009 |  |
| 60-70 | 0.75 | 0.71, 0.80 | <0.001 | <0.001 | 0.85 | 0.79, 0.90 | <0.001 | <0.001 |  |
| 70-80 | 0.64 | 0.59, 0.69 | <0.001 | <0.001 | 0.69 | 0.63, 0.75 | <0.001 | <0.001 |  |
| >80 | 0.35 | 0.31, 0.40 | <0.001 | <0.001 | 0.36 | 0.31, 0.41 | <0.001 | <0.001 |  |
| NIS SSS |  |  |  |  |  |  |  |  | 1.2 |
| I | — | — |  |  | — | — |  |  |  |
| II | 0.86 | 0.81, 0.92 | <0.001 | <0.001 | 1.01 | 0.94, 1.08 | 0.8 | >0.9 |  |
| III | 0.57 | 0.53, 0.61 | <0.001 | <0.001 | 0.79 | 0.73, 0.84 | <0.001 | <0.001 |  |
| Elixhauser Comorbidity Index | 0.98 | 0.97, 0.98 | <0.001 | <0.001 | 1.00 | 1.00, 1.01 | 0.031 | 0.2 | 1.1 |
| Hospital Location and Teaching Status |  |  |  |  |  |  |  |  | 1.2 |
|  | OR<sup>1</sup> | 95% CI<sup>1</sup> | p-value | q-value<sup>2</sup> | OR<sup>1</sup> | 95% CI<sup>1</sup> | p-value | q-value<sup>2</sup> | VIF<sup>1</sup> |
| *Urban, Non-Teach* | — | — |  |  | — | — |  |  |  |
| *Rural* | 1.76 | 1.11, 2.78 | **0.016** | 0.11 | 1.49 | 0.97, 2.31 | 0.071 | 0.4 |  |
| *Urban, Teach* | 0.63 | 0.55, 0.73 | **<0.001** | <0.001 | 0.84 | 0.71, 0.98 | **0.030** | 0.2 |  |
| **Hospital Region** |  |  |  |  |  |  |  |  | 1.2 |
| *Northeast* | — | — |  |  | — | — |  |  |  |
| *Midwest* | 1.09 | 0.94, 1.28 | 0.3 | >0.9 | 1.13 | 0.94, 1.35 | 0.2 | >0.9 |  |
| *South* | 1.06 | 0.92, 1.21 | 0.4 | >0.9 | 1.04 | 0.89, 1.21 | 0.6 | >0.9 |  |
| *West* | 1.15 | 1.00, 1.33 | 0.058 | 0.3 | 1.29 | 1.09, 1.52 | **0.002** | 0.022 |  |
| **Hospital Bed Size** |  |  |  |  |  |  |  |  | 1.1 |
| *Small* | — | — |  |  | — | — |  |  |  |
| *Medium* | 1.18 | 0.86, 1.61 | 0.3 | >0.9 | 1.15 | 0.88, 1.50 | 0.3 | >0.9 |  |
| *Large* | 1.13 | 0.83, 1.52 | 0.4 | >0.9 | 1.01 | 0.79, 1.29 | >0.9 | >0.9 |  |
| **Epoch** |  |  |  |  |  |  |  |  | 1.2 |
| *2001-2005* | — | — |  |  | — | — |  |  |  |
| *2006-2010* | 0.32 | 0.27, 0.38 | **<0.001** | <0.001 | 0.32 | 0.27, 0.38 | **<0.001** | <0.001 |  |
| *2011-2015* | 0.19 | 0.16, 0.23 | **<0.001** | <0.001 | 0.20 | 0.17, 0.23 | **<0.001** | <0.001 |  |
| *2016-2020* | 0.13 | 0.11, 0.15 | **<0.001** | <0.001 | 0.13 | 0.11, 0.16 | **<0.001** | <0.001 |  |

<sup>1</sup> OR = Odds Ratio, CI = Confidence Interval, GVIF = Generalized Variance Inflation Factor <sup>2</sup> Holm correction for multiple testing

**Supplemental Table 4:** Model parameters for multivariable regression for hospital mortality.

|  | Unadjusted Model |  |  |  | Adjusted Model |  |  |  |  |
| --- | --- | --- | --- | --- | --- | --- | --- | --- | --- |
|  | OR<sup>1</sup> | 95% CI<sup>1</sup> | p-value | q-value<sup>2</sup> | OR<sup>1</sup> | 95% CI<sup>1</sup> | p-value | q-value<sup>2</sup> | VIF<sup>1</sup> |
| **Microsurgery** | 0.91 | 0.85, 0.98 | **0.008** | 0.063 | 0.94 | 0.87, 1.02 | 0.13 | 0.8 | 1.2 |
| **Female Sex** | 0.98 | 0.92, 1.05 | 0.6 | >0.9 | 0.89 | 0.83, 0.96 | **0.001** | 0.012 | 1.0 |

|  | Unadjusted Model |  |  |  | Adjusted Model |  |  |  | VIF<sup>1</sup> |
| --- | --- | --- | --- | --- | --- | --- | --- | --- | --- |
|  | OR<sup>1</sup> | 95% CI<sup>1</sup> | p-value | q-value<sup>2</sup> | OR<sup>1</sup> | 95% CI<sup>1</sup> | p-value | q-value<sup>2</sup> |  |
| **Age Group** |  |  |  |  |  |  |  |  | 1.2 |
| *<50* | — | — |  |  | — | — |  |  |  |
| *50-60* | 1.28 | 1.17, 1.40 | **<0.001** | <0.001 | 1.19 | 1.09, 1.30 | **<0.001** | 0.002 |  |
| *60-70* | 1.67 | 1.53, 1.83 | **<0.001** | <0.001 | 1.46 | 1.32, 1.60 | **<0.001** | <0.001 |  |
| *70-80* | 2.49 | 2.25, 2.75 | **<0.001** | <0.001 | 2.06 | 1.85, 2.29 | **<0.001** | <0.001 |  |
| *>80* | 3.90 | 3.44, 4.42 | **<0.001** | <0.001 | 3.56 | 3.10, 4.09 | **<0.001** | <0.001 |  |
| **NIS SSS** |  |  |  |  |  |  |  |  | 1.1 |
| *I* | — | — |  |  | — | — |  |  |  |
| *II* | 2.94 | 2.70, 3.19 | **<0.001** | <0.001 | 2.98 | 2.73, 3.24 | **<0.001** | <0.001 |  |
| *III* | 4.35 | 4.00, 4.73 | **<0.001** | <0.001 | 4.51 | 4.13, 4.93 | **<0.001** | <0.001 |  |
| **Elixhauser Comorbidity Index** | 1.03 | 1.03, 1.03 | **<0.001** | <0.001 | 1.01 | 1.00, 1.01 | **0.034** | 0.2 | 1.2 |
| **Hospital Location and Teaching Status** |  |  |  |  |  |  |  |  | 1.1 |
| *Urban, Non-Teach* | — | — |  |  | — | — |  |  |  |
| *Rural* | 0.87 | 0.63, 1.21 | 0.4 | >0.9 | 0.96 | 0.70, 1.31 | 0.8 | >0.9 |  |
| *Urban, Teach* | 0.69 | 0.63, 0.77 | **<0.001** | <0.001 | 0.70 | 0.63, 0.78 | **<0.001** | <0.001 |  |
| **Hospital Region** |  |  |  |  |  |  |  |  | 1.2 |
| *Northeast* | — | — |  |  | — | — |  |  |  |
| *Midwest* | 0.83 | 0.74, 0.94 | **0.002** | 0.021 | 0.87 | 0.77, 0.98 | **0.027** | 0.2 |  |
| *South* | 0.95 | 0.86, 1.05 | 0.3 | >0.9 | 1.02 | 0.91, 1.13 | 0.8 | >0.9 |  |
| *West* | 1.09 | 0.98, 1.22 | 0.13 | 0.8 | 1.07 | 0.95, 1.21 | 0.3 | >0.9 |  |
| **Hospital Bed Size** |  |  |  |  |  |  |  |  | 1.1 |
| *Small* | — | — |  |  | — | — |  |  |  |
| *Medium* | 1.16 | 0.94, 1.44 | 0.2 | 0.8 | 1.12 | 0.90, 1.39 | 0.3 | >0.9 |  |
|  | OR<sup>1</sup> | 95% CI<sup>1</sup> | p-value | q-value<sup>2</sup> | OR<sup>1</sup> | 95% CI<sup>1</sup> | p-value | q-value<sup>2</sup> |  |
| *Large* | 1.11 | 0.91, 1.36 | 0.3 | >0.9 | 1.01 | 0.82, 1.24 | >0.9 | >0.9 |  |
| **Epoch** |  |  |  |  |  |  |  |  | 1.3 |
| *2001-2005* | — | — |  |  | — | — |  |  |  |
| *2006-2010* | 0.88 | 0.79, 0.97 | **0.014** | 0.10 | 0.75 | 0.67, 0.84 | **<0.001** | <0.001 |  |
| *2011-2015* | 0.77 | 0.69, 0.85 | **<0.001** | <0.001 | 0.53 | 0.48, 0.60 | **<0.001** | <0.001 |  |
| *2016-2020* | 0.88 | 0.80, 0.96 | **0.007** | 0.063 | 0.58 | 0.52, 0.65 | **<0.001** | <0.001 |  |

<sup>1</sup> OR = Odds Ratio, CI = Confidence Interval, VIF = Variance Inflation Factor <sup>2</sup> Holm correction for multiple testing

**Supplementary Table 5:**
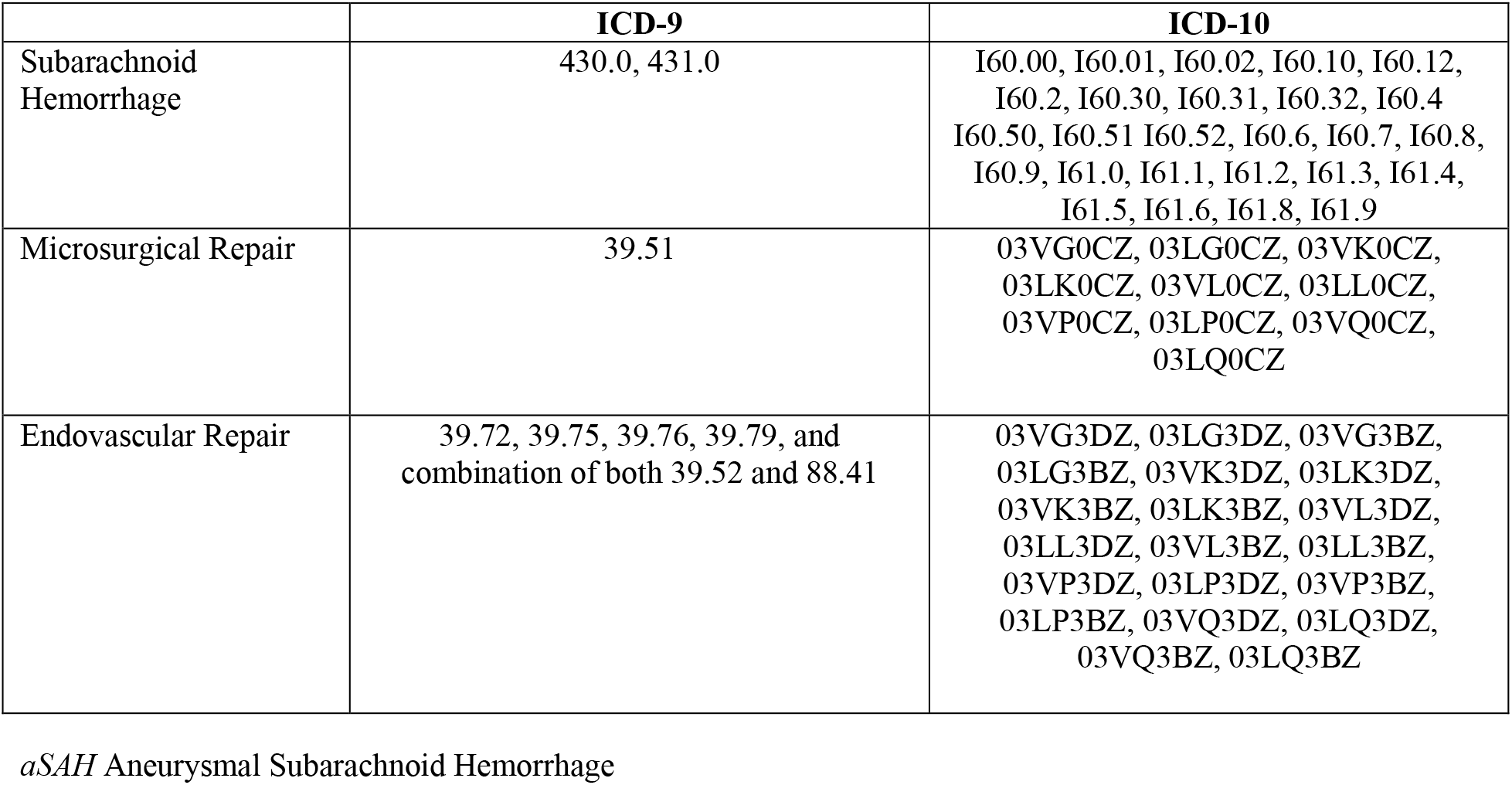
ICD-9-and-10 Codes for Identifying Aneurysmal Subarachnoid Hemorrhage Cases.

|  | ICD-9 | ICD-10 |
| --- | --- | --- |
| Subarachnoid Hemorrhage | 430.0, 431.0 | I60.00, I60.01, I60.02, I60.10, I60.12, I60.2, I60.30, I60.31, I60.32, I60.4 I60.50, I60.51 I60.52, I60.6, I60.7, I60.8, I60.9, I61.0, I61.1, I61.2, I61.3, I61.4, I61.5, I61.6, I61.8, I61.9 |
| Microsurgical Repair | 39.51 | 03VG0CZ, 03LG0CZ, 03VK0CZ, 03LK0CZ, 03VL0CZ, 03LL0CZ, 03VP0CZ, 03LP0CZ, 03VQ0CZ, 03LQ0CZ |
| Endovascular Repair | 39.72, 39.75, 39.76, 39.79, and combination of both 39.52 and 88.41 | 03VG3DZ, 03LG3DZ, 03VG3BZ, 03LG3BZ, 03VK3DZ, 03LK3DZ, 03VK3BZ, 03LK3BZ, 03VL3DZ, 03LL3DZ, 03VL3BZ, 03LL3BZ, 03VP3DZ, 03LP3DZ, 03VP3BZ, 03LP3BZ, 03VQ3DZ, 03LQ3DZ, 03VQ3BZ, 03LQ3BZ |

*aSAH* Aneurysmal Subarachnoid Hemorrhage

**Supplementary Table 6:** ICD 9-and-10 Codes for Determining the Severity of the Patient’s Presentation.

|  | ICD-9 | ICD-10 |
| --- | --- | --- |
| Coma | 780.01, 780.02, 780.03 780.09 | R40.0, R40.1, R40.20, R40.2A, R40.2420, R40.2421, R40.2422, R40.2423, R40.2424, R40.2430, R40.2431, R40.2432, R40.2433, R40.2434, R40.30, R40.4 |
| Hydrocephalus | 331.3, 331.4 | G91.0, G91.1, G91.9 |
| Hydrocephalus Treatment | 02.21, 02.22, 02.31, 02.32, 02.33, 02.34, 02.35, 02.39 | 009600Z, 009630Z, 009640Z, 001607B, 00160JB, 00160KB, 001637B, 00163JB, 00163KB, 001647B, 00164JB, 00164KB, 00963ZZ, 00964ZZ, 0016070, 0016071, 00160J0, 00160J1, 00160K0, 00160K1, 00160ZB, 0016370, 0016371, 00163J0, 00163J1, 00163K0, 00163K1, 00163ZB, 0016470, 0016471, 00164J0, 00164J1, 00164K0, 00164K1, 00164ZB, 0016072, 0016073, 00160J2, 00160J3, 00160K2, 00160K3, 0016372, 0016373, 00163J2, 00163J3, 00163K2, 00163K3, 0016472, 0016473, 00164J2, 00164J3, 00164K2, 0164K3, 0016074, 00160J4, 00160K4, 0016374, 00163J4, 00163K4, 0016474, 00164J4, 00164K4, 0W110J9, 0W110JB, 0016075, 0016076, 00160J5, 00160J6, 00160K5, 00160K6, 0016375, 0016376, 00163J5, 00163J6, 00163K5, 00163K6, 0016475, 0016476, 00164J5, 00164J6, 00164K5, 00164K6, 0W110JG, 0W110JJ, 0016077, |
|  |  | 00160J7, 00160K7, 0016377, 00163J7,<br>00163K7, 0016477, 00164J7,<br>00164K7, 0016078, 00160J8,<br>00160K8, 0016378, 00163J8,<br>00163K8, 0016478, 00164J8, 00164K8 |
| Cranial Nerve Deficits | 378.50, 378.51, 378.52, 378.53, 378.54,<br>378.55, 378.56, 379.40, 379.41, 379.42,<br>379.43, 379.45, 379.46, 379.49 | H49.00, H49.01, H49.02, H49.03,<br>H49.10, H49.11, H49.12, H49.13,<br>H49.20, H49.21, H49.22, H49.23,<br>H49.30, H49.31, H49.32, H49.33,<br>H49.40, H49.41, H49.42, H49.43,<br>H49.811, H49.812, H49.813, H49.819,<br>H49.881, H49.882, H49.883, H49.889,<br>H49.9, H57.00, H57.01, H57.02,<br>H57.03, H57.04, H57.051, H57.052,<br>H57.053, H57.059, H57.09 |
| Aphasia and Paresis | 438.0, 438.10, 438.11, 438.12, 438.13,<br>438.14, 438.19, 438.20, 438.21, 438.22,<br>438.30, 438.31, 438.32, 438.40, 438.41,<br>438.42, 438.50, 438.51, 438.52, 438.53,<br>438.6, 438.7, 438.8, 438.81, 438.82,<br>438.83, 438.84, 438.85, 438.89, 438.9 | All codes under I69 |
| Mechanical Ventilation | 960.5, 967.0, 967.1, 967.2 | 0BH17EZ, 0BH18EZ, 5A1935Z,<br>5A1945Z, 5A1955Z, 5A1935Z,<br>5A1945Z, 5A1955Z |

**Supplementary Table 7.** ICD 9-and-10 Codes for Identifying Admissions Outcomes.

|  | **ICD-9** | **ICD-10** |
| --- | --- | --- |
| Tracheostomy | 31.1, 31.21, 31.29 | 0B110F4, 0B113F4, 0B114F4 |
| Gastrostomy | 43.11, 43.19, 44.32 | 0DH63UZ, 0DH64UZ, 0D163J4,<br>0D16474, 0D164J4, 0D164K4,<br>0D164Z4 |

